# Right Heart Remodeling in End-Stage Pulmonary Arterial Hypertension and the Impact of Treatment Intensity

**DOI:** 10.1101/2025.02.10.25322038

**Authors:** Su-Gang Gong, Qi-Hang Zhang, Jia-Yi Zhang, Qian Zhang, Rui-Zhang, Hong-Ling Qiu, Ci-Jun Luo, Hui-Ting Li, Wen-Hui Wu, Ping Yuan, Jing He, Jian Xu, Jin-Ming Liu, Qin-Hua Zhao, Lan Wang

## Abstract

**Background:** Research on the limits of compensatory right heart remodeling and the effects of pulmonary artery hypertension (PAH) targeted therapies on these mechanisms is limited.

**Method:** Chest x-ray and echocardiographic data were collected from 143 deceased patients with PAH confirmed by right heart catheterization at their end-stage disease. Right heart remodeling was compared across different PAH treatment strategies.

**Results:** This study of 143 deceased PAH patients (49 ± 17 years, 74.1% female) characterized right heart remodeling at the time of death. Mean cardiothoracic ratio (CTR), right atrial area (RAA) and mid-cavity RV linear dimension (RVD) measured by echocardiography were 0.61±0.09, 27 cm² (median 27, IQR 21–38), and 4.97±0.97 cm, respectively, with extremes of 0.88, 102 cm², and 7.50 cm. Intensive therapy resulted in larger CTR (0.63±0.08 vs. 0.60±0.09, p=0.016), RAA (30 [24–40] vs. 25 [19–34] cm², p=0.020), and RVD (5.30±0.97 vs. 4.65±0.85 cm, p<0.001) compared with monotherapy. After adjusting for confounders, intensive therapy independently predicted increases in CTR (0.03, 95% CI 0.00-0.05, p=0.054), RAA (6.63 cm², 95% CI 1.46-11.80, p=0.013), and RVD (0.66 cm, 95% CI 0.34-0.98, p<0.001).

**Conclusion:** These findings suggest that more aggressive PAH treatment is associated with greater right heart remodeling, highlighting the complex relationship between therapeutic intervention and disease progression in PAH patients.

## Introduction

Pulmonary arterial hypertension (PAH) encompasses various conditions characterized by elevated pulmonary vascular resistance (PVR), including idiopathic, heritable, and PAH associated with connective tissue disease or congenital heart disease^1–3^. Persistent elevation in pulmonary artery pressure increases right ventricular afterload, triggering adaptive changes to maintain cardiac output^4^. Over time, chronic pressure overload eventually leads to maladaptive right ventricular (RV) remodeling, marked by myocardial cell proliferation and fibrosis. This structural and functional remodeling represents a critical turning point in PAH progression^5^. RV remodeling not only alters cardiac morphology but also impairs contractility, decreasing cardiac output and potentially leading to right heart failure, a major cause of mortality in PAH ^6^.

Significant advancements in drug therapies have markedly impacted right heart remodeling and improved the prognosis for patients with PAH^7^. Current treatments target four main signaling pathways: endothelin-1^8,9^, nitric oxide^10–12^, prostacyclin^13^, and the Activin/morphogenetic protein(BMP) pathway^14^. While monotherapy options exist, combination therapies^7^, including current triple and emerging quadruple drug regimens, have demonstrated greater efficacy. For those who remain unresponsive to these advanced therapies, lung transplantation remains a last resort.

Despite these therapeutic advances, disease progression, irreversible RV remodeling, and death still occur. Currently, limited research explores the limits of RV remodeling compensation and the impact of targeted therapies on these compensatory mechanisms. Therefore, further investigation is warranted to determine how these pharmacological strategies affect the limits of RV adaptation and to elucidate whether these limits vary among PAH patients.

## Methods

### Study Design and Patient Selection

This retrospective cohort study was conducted in department of pulmonary circulation, Shanghai Pulmonary Hospital. All patients diagnosed with PAH in our department undergo regular follow-up, and for those whose deaths are confirmed during this follow-up period, we gather their information. We collected clinical data at baseline and the last visit data of these patients, encompassing demographic characteristics, laboratory test results, echocardiographic findings, chest radiographs and hemodynamic data. Our study adhered to the principles of the Declaration of Helsinki and received approval from the ethics committee of Shanghai Pulmonary Hospital (approval number: K16-293).

Patients had to meet the following criteria to be included in the study: (i) diagnosis between January 1, 2013, and June 30, 2024; (ii) confirmed diagnosis of Group 1 PAH, defined as mean pulmonary arterial pressure (mPAP) > 20 mmHg, pulmonary artery wedge pressure(PAWP) < 15mmHg and pulmonary vascular resistance(PVR) > 2WU by right heart catheterization (RHC) at the time of diagnosis; (iii) visit data available within one year before death; (iv) death confirmed by telephone or visit. Patients who did not meet the diagnostic criteria for PAH and those lacking relevant visit data within one year before death were excluded.

### Clinical, Functional and Hemodynamic Characteristics

A comprehensive evaluation of clinical data was performed, including demographic information such as age, sex, as well as medical and family histories. The hemodynamic data were collected, including mPAP, PAWP and right atrial pressure (RAP). The cardiac index (CI) was determined by measuring cardiac output (CO) with the standard thermodilution technique and divided by body surface area. PVR was calculated using the formula: (mPAP-PAWP)/CO^15^.

### Cardiothoracic ratio and Echocardiography assessment

The cardiothoracic ratio (CTR) on a chest X-ray is determined by comparing the width of the heart to the width of the chest ^16^. All patients underwent CTR measurements at both baseline and during their last visit All echocardiographic data were acquired using commercially available equipment (Vivid 7, GE Healthcare) in standard views. Measurements were obtained from the mean of three consecutive beats based on the American Society of Echocardiography Guidelines^17^. The echo parameters and derived assessments that we focused on common and widely available for daily clinical practice, including RA area, mid-cavity RV linear dimension(RVD), left ventricular end diastolic diameter (LVEDD), left ventricular ejection fraction (LVEF), left ventricular end-diastolic eccentricity index (LV-EId), pulmonary arterial systolic pressure (PASP), tricuspid annular plane systolic excursion (TAPSE), and presence of pericardial effusion.

### Statistical Analyses

All statistical analyses were performed using SPSS version 20.0 software (IBM Corp., Armonk, NY, USA) and GraphPad Prism version 8 (GraphPad Software Inc., La Jolla, CA, USA). Categorical data are presented as numbers and percentages, while continuous data are presented as either mean ± SD or median with interquartile range (IQR), depending on the data distribution. Continuous data were compared using Welch Two Sample t-test or one-way ANOVA for parametric data, and Wilcoxon signed-rank/Kruskal-Wallis rank sum test for nonparametric data. Linear regression analysis is used to describe the relationship between intensity of PAH therapy and the extent of heart remodeling. Multiple models were constructed, each adjusting for a different set of covariates to provide a nuanced understanding of how these covariates influence the observed association. A p-value of <0.05 was considered significant for all statistical tests.

## Results

### Characteristics of Study Population

A total of 143 deceased PAH patients met the inclusion and exclusion criteria. Demographic, classification, hemodynamic, treatment, and cardiac remodeling data of the study population are presented in **Table 1**. The average age was 49 ± 17 years, with 37 (25.9%) males and 106 (74.1%) females. Time from symptom onset to death was 6.3 ± 4.5 years, and time from diagnosis to death was 3.4 ± 3.6 years. PAH classifications were idiopathic (63, 44.1%), heritable (4, 2.8%), connective tissue disease (51, 35.7%), portal hypertension (2, 1.4%), congenital heart disease (20, 14.0%), and PVOD/PCH (3, 2.1%).

**Table 1:** Clinical Characteristics of Deceased Patients with PAH.

| Characteristic | N = 143<sup>1</sup> |
| --- | --- |
| Age, yr | 49 ± 17 |
| Sex |  |
| Male | 37 (25.9%) |
| Female | 106 (74.1%) |
| Symptom Onset Time<sup>2</sup>, yr | 6.3 ± 4.5 |
| Diagnosis Time<sup>3</sup>, yr | 3.4 ± 3.6 |
| PAH Classifications |  |
| Idiopathic | 63 (44.1%) |
| Heritable | 4 (2.8%) |
| Connective tissue disease | 51 (35.7%) |
| Portal hypertension | 2 (1.4%) |
| Congenital heart disease | 20 (14.0%) |
| PVOD/PCH | 3 (2.1%) |
| Baseline RHC<sup>4</sup> |  |
| PAP<sub>m</sub>, mmHg | 57 ± 16 |
| PAWP, mmHg | 7.1 ± 4.4 |
| RAP, mmHg | 6.1 ± 5.0 |
| CO, L/min | 3.08 ± 2.23 |
| CI, L/min/m<sup>2</sup> | 2.60 ± 1.11 |
| PVR, Wood units | 13 ± 7 |
| SVO<sub>2</sub>, % | 58 ± 14 |
| PAH therapy |  |
| Mono-therapy | 20 (14.0%) |
| Double-therapy | 56 (39.2%) |
| Triple-therapy | 67 (46.9%) |
| Cause of death |  |
| Sudden death | 41 (28.7%) |
| Right heart failure | 89 (62.2%) |
| Other<sup>5</sup> | 13 (9.1%) |

<sup>1</sup>Mean ± SD; n (%)
<sup>2</sup>Time from first reported PAH symptom to death.
<sup>3</sup>Time from right heart catheterization (RHC) confirming the diagnosis of PAH to death.
<sup>4</sup>Data presented represent the patients' status at the time of death, except for RHC data, which were collected at baseline
<sup>5</sup>Other causes included infection (n=6), hemoptysis (n=3), cerebral hemorrhage (n=2), transplant-related death (n=1), and colonoscopy complications (n=1).
*Definition of abbreviations:* CI: Cardiac Index; CO: Cardiac Output; MPAP: Mean Pulmonary Artery Pressure; PAH: Pulmonary Arterial Hypertension; PASP: Pulmonary Artery Systolic Pressure; PAWP: Pulmonary Artery Wedge Pressure; PCH: Pulmonary Capillary Haemangiomatosis; PVR: Pulmonary Vascular Resistance; PVOD: Pulmonary Veno-Occlusive Disease; RAP: Right Atrial Pressure; RHC: Right Heart Catheterization; SvO<sub>2</sub>: Mixed Venous Oxygen Saturation.

At diagnosis, right heart catheterization demonstrated mPAP of 57 ± 16 mmHg, PAWP of 7.1 ± 4.4 mmHg, RAP of 6.1 ± 5.0 mmHg, CO of 3.08 ± 2.23 L/min, CI of 2.60 ± 1.11 l/min/m², PVR of 13 ± 7 Wood units, and a mixed venous oxygen saturation (SvO₂) of 58 ± 14%. With respect to PAH-specific treatments, 20 (14.0%) received monotherapy (14 on phosphodiesterase type 5 [PDE5] inhibitors, 6 on endothelin receptor antagonists [ERAs]), 56 (39.2%) received double therapy (all on PDE5 inhibitors plus ERAs), and 67 (46.9%) were treated with triple therapy (ERAs, PDE5 inhibitors, and prostacyclin analogues). The primary cause of death was right heart failure in 89 (62.2%) patients. Sudden death accounted for 41 (28.7%) deaths. Other causes of death included infection (n=6), hemoptysis (n=3), cerebral hemorrhage (n=2), transplant-related death (n=1), and colonoscopy complications (n=1).

### Right Heart Remodeling at the Time of Death

At death, chest radiography showed a CTR of 0.61 ± 0.09. Echocardiography revealed RAA of 27 cm² (median 27, IQR 21–38), RVD of 4.97 ± 0.97 cm, LVEDD of 3.40 cm (median 3.40, IQR 3.00–4.00), and LV-EID of 1.56 (median 1.56, IQR 1.34–1.75) **(Table 2).** Notably, the extreme limits of compensatory remodeling in end-stage disease included a CTR as high as 0.88, an RAA of 102 cm², and an RVD of 7.50 cm. Figure 1 illustrates the frequency distribution of right ventricular remodeling parameters at death.

**Figure 1.**
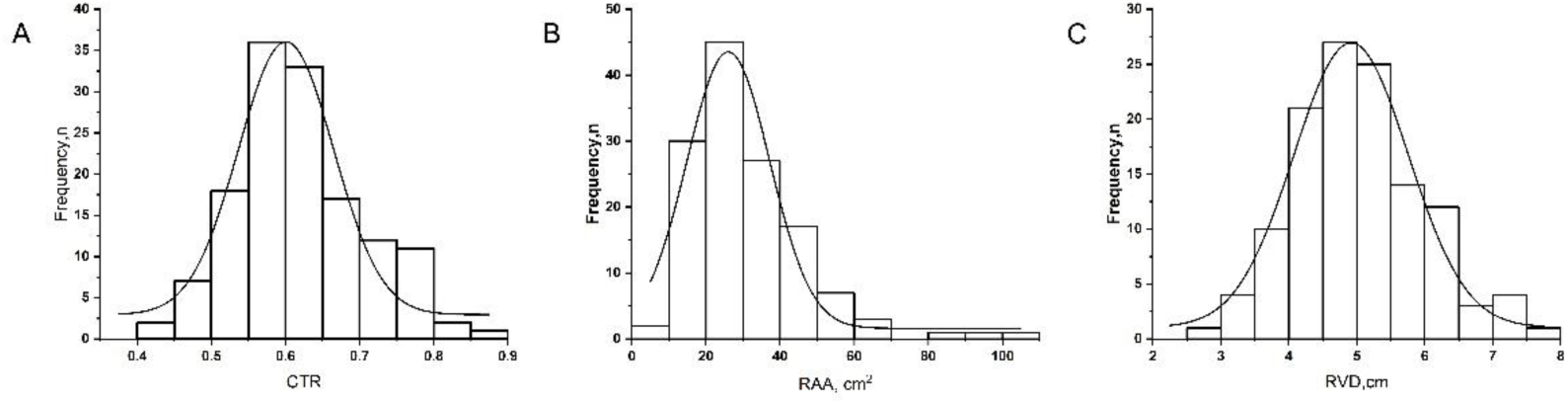
Frequency distribution of right ventricular remodeling parameters at the time of death in patients with pulmonary arterial hypertension. The histograms show the distribution of (A) CTR (cardiothoracic ratio measured from chest X-ray), (B) RAA (Right Atrial Area measured by echocardiography, cm²), and (C) RVD (Right Ventricular Diameter measured by echocardiography, cm). The superimposed curves represent unimodal probability density functions fitted to the respective histograms.

**Table 2.**
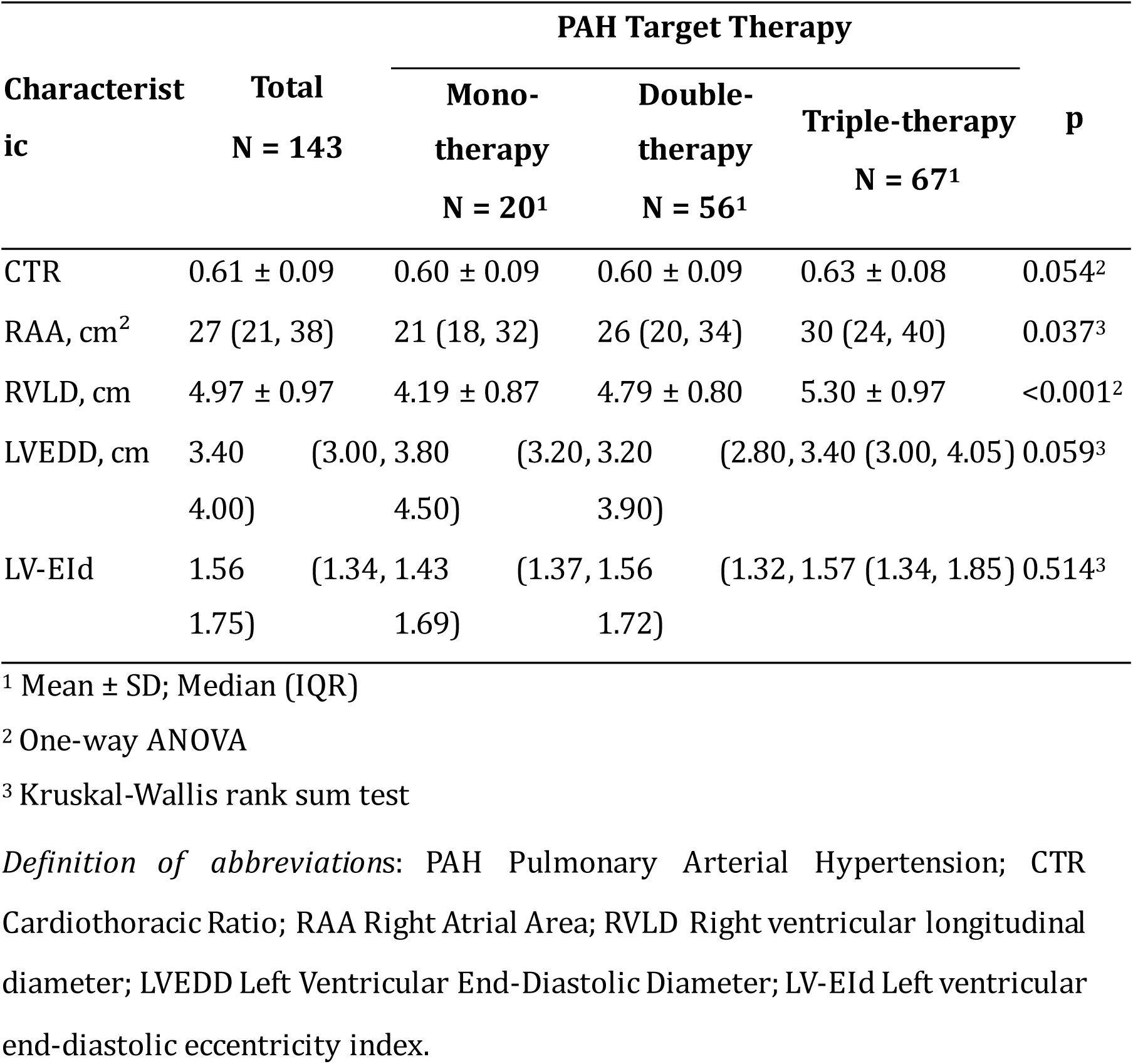
Compensatory Cardiac Remodeling in PAH Patients Under Different Treatment Strategies.

Before death, varying treatment strategies were associated with different degrees of right heart remodeling **(Table 2)**. Monotherapy (20 patients) resulted in a CTR of 0.60 ± 0.09, RAA of 21 cm² (median 18, IQR 18–32), and RVD of 4.19 ± 0.87 cm; double therapy (56 patients) showed a CTR of 0.60 ± 0.09, RAA of 26 cm² (median 20, IQR 20–34), and RVDof 4.79 ± 0.80 cm; and triple therapy (67 patients) led to a CTR of 0.63 ± 0.08, RAA of 30 cm² (median 24, IQR 24–40), and RVD of 5.30 ± 0.97 cm. Statistical analysis indicated significant differences between treatment groups for RAA (p=0.037) and RVD (p<0.001), and a near-significant difference for CTR (p=0.054). LVEDD and LV-EID remained relatively consistent across the different treatment approaches.

Compared with those who were insufficiently treated, patients receiving intensive combination therapy had a significantly larger RVD (5.30 ± 0.97 cm vs. 4.65 ± 0.85 cm, p<0.001) and RAA (30 [24–40] cm² vs. 25 [19–34] cm², p=0.020) **(Table 3**, **Figure 2)**. However, no significant differences were observed in LVEDD or LV-EID. The CTR was slightly higher under intensive combination therapy (0.63 ± 0.08 vs. 0.60 ± 0.09, p=0.016).

**Figure 2.**
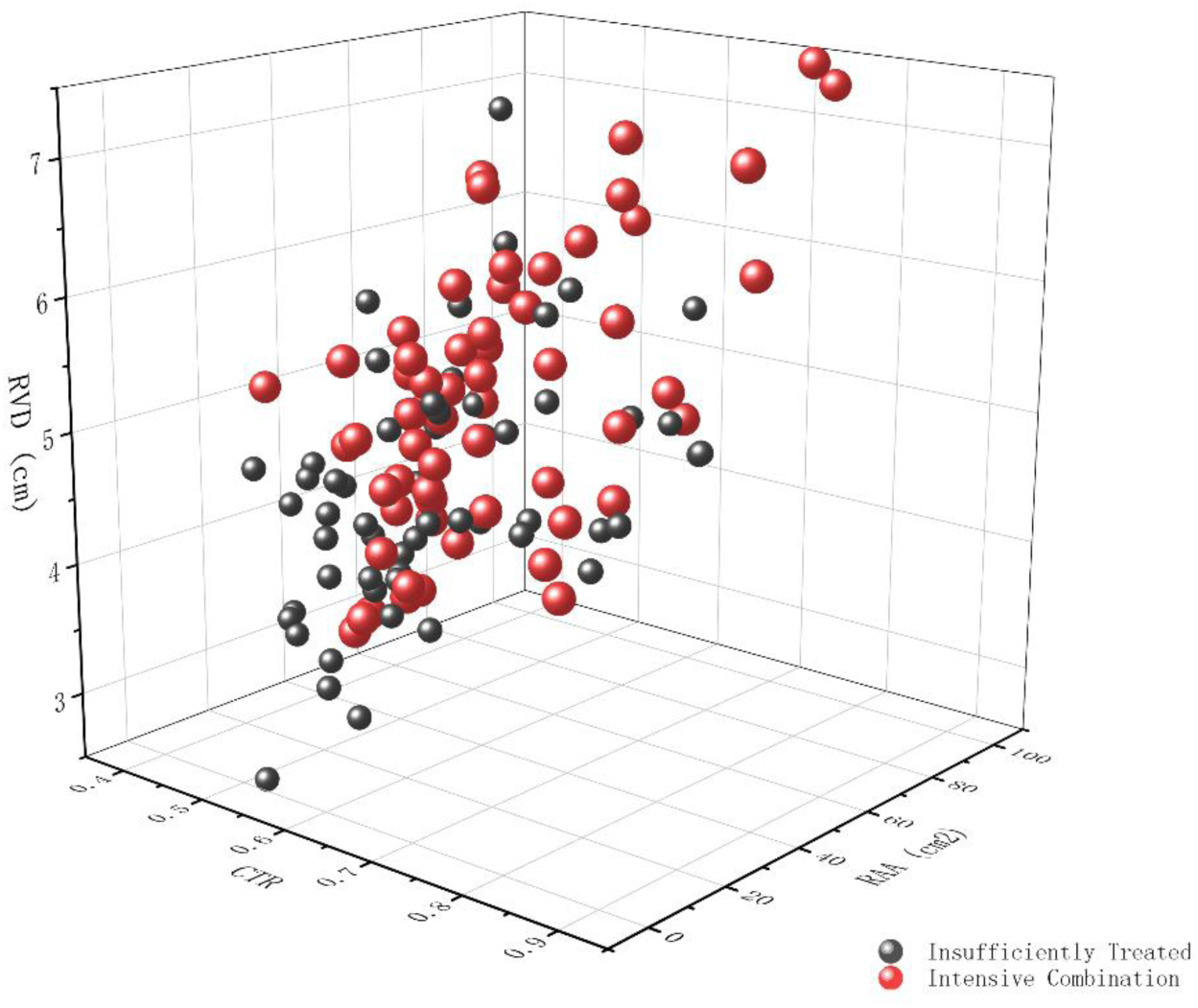
Three-dimensional scatter plot demonstrating the impact of treatment intensity on right ventricular remodeling. The plot depicts the relationship between cardiothoracic ratio (CTR) measured from chest X-ray, right ventricular diameter (RVD, cm), and right atrial area (RAA, cm²) measured by echocardiography, in patients receiving either insufficient treatment (dark grey spheres) or intensive combination therapy (red spheres).

**Table 3.** Compensatory Cardiac Remodeling in PAH Comparing Insufficient and Intensive Therapy Approaches.

| Characteristic | PAH Target Therapy |  | p |
| --- | --- | --- | --- |
|  | Insufficiently Treated<br>N = 76<sup>1</sup> | Intensive Combination N =<br>67<sup>1</sup> |  |
| CTR | 0.60 ± 0.09 | 0.63 ± 0.08 | 0.016<sup>2</sup> |
| RAA, cm<sup>2</sup> | 25 (19, 34) | 30 (24, 40) | 0.020<sup>3</sup> |
| RVLD, cm | 4.65 ± 0.85 | 5.30 ± 0.97 | <0.001<sup>2</sup> |
| LVEDD, cm | 3.35 (2.93, 4.00) | 3.40 (3.00, 4.05) | 0.845<sup>3</sup> |
| LV-EId | 1.55 (1.32, 1.71) | 1.57 (1.34, 1.85) | 0.299<sup>3</sup> |

<sup>1</sup>Mean ± SD; Median (IQR)
<sup>2</sup>Welch Two Sample t-test
<sup>3</sup>Wilcoxon rank sum test
*Definition of abbreviations:* PAH Pulmonary Arterial Hypertension; CTR Cardiothoracic Ratio; RAA Right Atrial Area; RVLD Right ventricular longitudinal diameter; LVEDD Left Ventricular End-Diastolic Diameter; LV-EId Left ventricular end-diastolic eccentricity index.

### Impact of PAH Therapy on Right Ventricular Remodeling Limits

Linear regression analysis demonstrated a clear association between the intensity of PAH therapy and the extent of right ventricular remodeling, as detailed in Table 4. Patients receiving intensive combination therapy experienced significantly greater increases in CTR, RAA, and RVD compared to those receiving insufficient therapy. The analysis revealed that intensive therapy led to a 0.04 increase in CTR (95% CI 0.01-0.06, p=0.016), a 6.12 cm² increase in RAA (95% CI 0.87-11.36, p=0.024), and a 0.65 cm increase in RVD (95% CI 0.33-0.98, p<0.001).

**Table 4:** Association between PAH Therapy Intensity and Compensatory Cardiac Remodeling (Linear regression)

| Characteristic | Unadjusted |  |  | Model I |  |  | Model II |  |  | Model III |  |  |
| --- | --- | --- | --- | --- | --- | --- | --- | --- | --- | --- | --- | --- |
|  | Beta | 95% CI | p | Beta | 95% CI | p-value | Beta | 95% CI | p | Beta | 95% CI | p |
| **CTR** |  |  |  |  |  |  |  |  |  |  |  |  |
| Insufficient Therapy | Ref |  |  | Ref |  |  | Ref |  |  | Ref |  |  |
| Intensive Therapy | 0.04 | 0.01, 0.06 | 0.016 | 0.03 | 0.00, 0.06 | 0.016 | 0.03 | 0.00, 0.06 | 0.051 | 0.03 | 0.00, 0.05 | 0.054 |
| **RAA** |  |  |  |  |  |  |  |  |  |  |  |  |
| Insufficient Therapy | Ref |  |  | Ref |  |  | Ref |  |  | Ref |  |  |
| Intensive Therapy | 6.12 | 0.87, 11.36 | 0.024 | 5.67 | 0.40,10.95 | 0.037 | 6.85 | 1.62, 12.08 | 0.011 | 6.63 | 1.46, 11.80 | 0.013 |
| **RVLD** |  |  |  |  |  |  |  |  |  |  |  |  |
| Insufficient Therapy | Ref |  |  | Ref |  |  | Ref |  |  | Ref |  |  |
| Intensive Therapy | 0.65 | 0.33,0.98 | <0.001 | 0.63 | 0.31,0.95 | <0.001 | 0.67 | 0.35, 0.99 | <0.001 | 0.66 | 0.34, 0.98 | <0.001 |

Model I: adjusted for age and gender;
Model II: adjusted for Model I+ PAH Classifications;
Model III: adjusted for Model II+ PAH Classifications+ Cause of death.
Definition of abbreviations: PAH Pulmonary Arterial Hypertension; CTR Cardiothoracic Ratio; RAA Right Atrial Area; RVLD Right ventricular longitudinal diameter; CI Confidence Interval.

Adjusting for age and sex (Model I) maintained the statistical significance of these findings, the increase in CTR was 0.03 (95% CI 0.00-0.06, p=0.016), the increase in RAA was 5.67 cm² (95% CI 0.40-10.95, p=0.037), and the increase in RVD was 0.63 cm (95% CI 0.31-0.95, p<0.001). Further adjusting for PAH classifications (Model II) also demonstrated significant increases with intensive therapy for CTR (0.03, 95% CI 0.00-0.06, p=0.051), RAA (6.85 cm², 95% CI 1.62-12.08, p=0.011), and RVD (0.67 cm, 95% CI 0.35-0.99, p<0.001). Finally, even after adjusting for cause of death in addition to age, sex, and PAH classifications (Model III), intensive therapy remained significantly associated with increased CTR (0.03, 95% CI 0.00-0.05, p=0.054), RAA (6.63 cm², 95% CI 1.46-11.80, p=0.013), and RVD (0.66 cm, 95% CI 0.34-0.98, p<0.001).

## Discussion

The present study demonstrates that patients with PAH exhibit varying degrees of right heart remodeling in the terminal stage, with extreme cases showing CTR up to 0.88, RAA up to 102 cm², and RVD up to 7.50 cm. Intensive combination therapy (triple therapy) was significantly associated with more pronounced right heart remodeling compared to monotherapy or double therapy. Specifically, patients receiving triple therapy had larger CTRs, RAAs, and RVDs, as evidenced by both chest X-rays and echocardiography. Linear regression analysis confirmed that the intensity of PAH therapy independently correlated with increased right ventricular remodeling, regardless of age, sex, PAH classification and cause of death.

PAH is a progressive condition characterized by escalating pulmonary vascular resistance, which places an increased afterload on the right heart^2,18,19^. In response, the right ventricle initially compensates by thickening its walls and enhancing contractility^19,20^. Over time, however, sustained pressure overload can lead to significant right heart enlargement, a process intimately linked to patient survival^21–24^. Although extreme cases of right heart remodeling in PAH are rarely documented, our findings indicate a remarkable capacity for structural adaptation, with right atrial areas reaching five to six times the upper limit of normal and right ventricular diameters exceeding twice the normal threshold^25^. This degree of remodeling is shaped by an intricate interplay of factors, including neurohormonal activation, coronary perfusion, myocardial metabolism, disease onset rate, underlying etiology, and emerging genetic or epigenetic influences^20,26,27^. Notably, our findings demonstrate that therapeutic strategies are another significant clinical factor influencing right heart remodeling. While cardiac dimensions are influenced by various confounding factors such as age^28^, sex^29^, PAH classification^30^, and cause of death^31^, our analysis demonstrates that even after adjusting for these factors, treatment intensity remains significantly associated with right heart remodeling.

PAH-targeted therapies have been shown to influence right heart remodeling^32–34^. Novel agents like sotatercept improve right ventricular structure by reducing pulmonary vascular resistance^35^, thereby prolonging the clinical course of PAH. Focusing on the concept of cardiac compensatory limits, our study elucidates the complex relationship between PAH-targeted therapy and right heart remodeling. We found a positive correlation between right heart size and treatment intensity, with significantly greater remodeling observed in patients receiving triple therapy before death compared to those without adequate treatment. Furthermore, the extent of right heart remodeling demonstrated a linear relationship with the duration of PAH management (data not shown). These results suggest that more comprehensive pharmacologic treatment allows for greater cardiac compensation, effectively extending right heart adaptation closer to its functional limits.

Right heart remodeling in PAH presents a complex duality. While the heart’s initial remodeling response to increased pulmonary artery pressure is beneficial, enabling it to compensate and prolong survival^19,36^, especially with therapeutic intervention, this adaptive process eventually becomes detrimental. Prolonged remodeling leads to significant right heart enlargement, a key predictor of poor prognosis^37^. This creates a critical challenge: While early intervention aims to reverse remodeling, the focus for advanced disease shifts to slowing its progression and maximizing the heart’s compensatory capacity.

The emergence of “super-remodelers”—patients exhibiting extreme right heart enlargement despite prolonged PAH-targeted therapy—poses a significant challenge in lung transplantation decisions. While previous research suggests comparable outcomes between bilateral lung transplantation (BLT) and heart-lung transplantation (HLT)^38,39^, the decision-making process becomes more complex for these individuals. Although improved survival rates with PAH medications may be reducing the overall need for lung transplants, the rise of extreme right heart remodeling necessitates a closer look at optimal surgical strategies for this specific patient population. Further research focusing on the potential reversibility of this severe remodeling post-transplant is crucial for refining treatment approaches and determining whether BLT or HLT offers the best long-term outcomes.

This study has several limitations. First, its retrospective design means baseline data were collected at our center upon diagnosis. Many patients had received prior treatment at other facilities, precluding a truly treatment-naï ve assessment of initial cardiac structure. This inability to account for pre-referral treatment history limits our understanding of the true baseline right heart characteristics. Second, we lacked data on treatment changes over the disease course, hindering analysis of dynamic interactions between evolving treatment strategies and right heart remodeling progression. This prevented us from evaluating the impact of treatment modifications on remodeling over time. Third, cardiac remodeling at the time of death was assessed using chest radiography and echocardiography^40–42^. While these modalities are readily available and accepted methods for evaluating right heart structure, the absence of MRI data limited our ability to fully characterize end-stage remodeling, especially in cases with extreme enlargement.

## Conclusion

Right heart remodeling in PAH progresses throughout the disease course, culminating in significant changes in the terminal stage. PAH-targeted therapies influence the extent of this cardiac remodeling, effectively pushing the limits of compensatory adaptation. This influence is independent of age, sex, PAH classification, and cause of death. While these therapies enhance the heart’s compensatory capacity, allowing it to reach structural extremes, this phenomenon presents a complex duality. The emergence of “super-remodelers,” patients exhibiting extreme right heart enlargement despite prolonged PAH-targeted therapy, underscores the challenges in managing advanced PAH, particularly regarding decisions surrounding lung transplantation.

## Data Availability

all data is available

## Contributors

All authors participated in the design of the study and/or patient enrolment, and meet criteria for authorship. S.G.G, Q.H.Z. and L.W. contributed to the study design, study conduct and supervision, scientific overview, data analysis, and editing of the manuscript. Q.H.Z., S.G.G., J.Y.Z., Q.H.Z., and L.W. draft and edit the original manuscript. Q.H.Z., J.Y.Z., Q.Z., R.Z., C.J.L., H.L.Q., W.H.W., H.T.L., J.H., P.Y., J.M.L., S.G.G., and L.W. generated the data. Q.H.Z., S.G.G., and L.W. analyzed the data. Q.H.Z., R.Z., S.G.G., and L.W. interpreted the data. Q.H.Z., S.G.G., C.J.L., H.L.Q., W.H.W., H.T.L., J.H., P.Y., J.X., J.M.L., S.G.G., and L.W. contributed to patient enrolment and clinical interpretation. All authors have reviewed the manuscript and approved the final version for submission.

## Reference

1. Randall PA, Heitzman ER, Bull MJ, et al. Pulmonary arterial hypertension: a contemporary review. Radiographics 1989;9:905–927. doi: 10.1148/radiographics.9.5.2678297

2. Hassoun PM. Pulmonary Arterial Hypertension. N Engl J Med 2021;385:2361–2376. doi: 10.1056/NEJMra2000348

3. Kovacs G, Bartolome S, Denton CP, et al. Definition, classification and diagnosis of pulmonary hypertension. Eur Respir J 2024;64. doi: 10.1183/13993003.01324-2024

4. Dini FL, Pugliese NR, Ameri P, et al. Right ventricular failure in left heart disease: from pathophysiology to clinical manifestations and prognosis. Heart Fail Rev 2023;28:757–766. doi: 10.1007/s10741-022-10282-2

5. Werbner B, Tavakoli-Rouzbehani OM, Fatahian AN, Boudina S. The dynamic interplay between cardiac mitochondrial health and myocardial structural remodeling in metabolic heart disease, aging, and heart failure. J Cardiovasc Aging 2023;3. doi: 10.20517/jca.2022.42

6. Naeije R, Richter MJ, Rubin LJ. The physiological basis of pulmonary arterial hypertension. Eur Respir J 2022;59. doi: 10.1183/13993003.02334-2021

7. Chin KM, Gaine SP, Gerges C, et al. Treatment algorithm for pulmonary arterial hypertension. Eur Respir J 2024;64. doi: 10.1183/13993003.01325-2024

8. Pulido T, Adzerikho I, Channick RN, et al. Macitentan and morbidity and mortality in pulmonary arterial hypertension. N Engl J Med 2013;369:809–818. doi: 10.1056/NEJMoa1213917

9. Galie N, Olschewski H, Oudiz RJ, et al. Ambrisentan for the treatment of pulmonary arterial hypertension: results of the ambrisentan in pulmonary arterial hypertension, randomized, double-blind, placebo-controlled, multicenter, efficacy (ARIES) study 1 and 2. Circulation 2008;117:3010–3019. doi: 10.1161/CIRCULATIONAHA.107.742510

10. Galie N, Brundage BH, Ghofrani HA, et al. Tadalafil therapy for pulmonary arterial hypertension. Circulation 2009;119:2894–2903. doi: 10.1161/CIRCULATIONAHA.108.839274

11. Sastry BK, Narasimhan C, Reddy NK, Raju BS. Clinical efficacy of sildenafil in primary pulmonary hypertension: a randomized, placebo-controlled, double-blind, crossover study. J Am Coll Cardiol 2004;43:1149–1153. doi: 10.1016/j.jacc.2003.10.056

12. Ghofrani HA, Galie N, Grimminger F, et al. Riociguat for the treatment of pulmonary arterial hypertension. N Engl J Med 2013;369:330–340. doi: 10.1056/NEJMoa1209655

13. Simonneau G, Barst RJ, Galie N, et al. Continuous subcutaneous infusion of treprostinil, a prostacyclin analogue, in patients with pulmonary arterial hypertension: a double -blind, randomized, placebo-controlled trial. Am J Respir Crit Care Med 2002;165:800–804. doi: 10.1164/ajrccm.165.6.2106079

14. Hoeper MM, Badesch DB, Ghofrani HA, et al. Phase 3 Trial of Sotatercept for Treatment of Pulmonary Arterial Hypertension. N Engl J Med 2023;388:1478–1490. doi: 10.1056/NEJMoa2213558

15. Humbert M, Kovacs G, Hoeper MM, et al. 2022 ESC/ERS Guidelines for the diagnosis and treatment of pulmonary hypertension. Eur Respir J 2023;61. doi: 10.1183/13993003.00879-2022

16. Truszkiewicz K, Poreba R, Gac P. Radiological Cardiothoracic Ratio in Evidence-Based Medicine. J Clin Med 2021;10. doi: 10.3390/jcm10092016

17. Rudski LG, Lai WW, Afilalo J, et al. Guidelines for the echocardiographic assessment of the right heart in adults: a report from the American Society of Echocardiography endorsed by the European Association of Echocardiography, a registered branch of the European Society of Cardiology, and the Canadian Society of Echocardiography. J Am Soc Echocardiogr 2010;23:685–713; quiz 786-688. doi: 10.1016/j.echo.2010.05.010

18. Simonneau G, Montani D, Celermajer DS, et al. Haemodynamic definitions and updated clinical classification of pulmonary hypertension. Eur Respir J 2019;53. doi: 10.1183/13993003.01913-2018

19. Vonk Noordegraaf A, Westerhof BE, Westerhof N. The Relationship Between the Right Ventricle and its Load in Pulmonary Hypertension. J Am Coll Cardiol 2017;69:236–243. doi: 10.1016/j.jacc.2016.10.047

20. Vonk-Noordegraaf A, Haddad F, Chin KM, et al. Right heart adaptation to pulmonary arterial hypertension: physiology and pathobiology. J Am Coll Cardiol 2013;62:D22–33. doi: 10.1016/j.jacc.2013.10.027

21. Raymond RJ, Hinderliter AL, Willis PW, et al. Echocardiographic predictors of adverse outcomes in primary pulmonary hypertension. J Am Coll Cardiol 2002;39:1214–1219. doi: 10.1016/s0735-1097(02)01744-8

22. van Wolferen SA, Marcus JT, Boonstra A, et al. Prognostic value of right ventricular mass, volume, and function in idiopathic pulmonary arterial hypertension. Eur Heart J 2007;28:1250–1257. doi: 10.1093/eurheartj/ehl477

23. Vonk Noordegraaf A, Chin KM, Haddad F, et al. Pathophysiology of the right ventricle and of the pulmonary circulation in pulmonary hypertension: an update. Eur Respir J 2019;53. doi: 10.1183/13993003.01900-2018

24. Badagliacca R, Poscia R, Pezzuto B, et al. Prognostic relevance of right heart reverse remodeling in idiopathic pulmonary arterial hypertension. J Heart Lung Transplant 2017. doi: 10.1016/j.healun.2017.09.026

25. Soulat-Dufour L, Addetia K, Miyoshi T, et al. Normal Values of Right Atrial Size and Function According to Age, Sex, and Ethnicity: Results of the World Alliance Societies of Echocardiography Study. J Am Soc Echocardiogr 2021;34:286–300. doi: 10.1016/j.echo.2020.11.004

26. Voelkel NF, Quaife RA, Leinwand LA, et al. Right ventricular function and failure: report of a National Heart, Lung, and Blood Institute working group on cellular and molecular mechanisms of right heart failure. Circulation 2006;114:1883–1891. doi: 10.1161/CIRCULATIONAHA.106.632208

27. Hemnes AR, Celermajer DS, D’Alto M, et al. Pathophysiology of the right ventricle and its pulmonary vascular interaction. Eur Respir J 2024;64. doi: 10.1183/13993003.01321-2024

28. Henein M, Waldenstrom A, Morner S, Lindqvist P. The normal impact of age and gender on right heart structure and function. Echocardiography 2014;31:5–11. doi: 10.1111/echo.12289

29. Westaby JD, Zullo E, Bicalho LM, Anderson RH, Sheppard MN. Effect of sex, age and body measurements on heart weight, atrial, ventricular, valvular and sub -epicardial fat measurements of the normal heart. Cardiovasc Pathol 2023;63:107508. doi: 10.1016/j.carpath.2022.107508

30. Ascha M, Renapurkar RD, Tonelli AR. A review of imaging modalities in pulmonary hypertension. Ann Thorac Med 2017;12:61–73. doi: 10.4103/1817-1737.203742

31. Tonelli AR, Arelli V, Minai OA, et al. Causes and circumstances of death in pulmonary arterial hypertension. Am J Respir Crit Care Med 2013;188:365–369. doi: 10.1164/rccm.201209-1640OC

32. Vonk Noordegraaf A, Channick R, Cottreel E, et al. The REPAIR Study: Effects of Macitentan on RV Structure and Function in Pulmonary Arterial Hypertension. JACC Cardiovasc Imaging 2022;15:240–253. doi: 10.1016/j.jcmg.2021.07.027

33. Zhao QH, Chen J, Chen FD, et al. Evaluating the efficacy and safety of oral triple sequential combination therapy for treating patients with pulmonary arterial hypertension: A multicenter retrospective study. Pulm Circ 2024;14:e12351. doi: 10.1002/pul2.12351

34. D’Alto M, Badagliacca R, Argiento P, et al. Risk Reduction and Right Heart Reverse Remodeling by Upfront Triple Combination Therapy in Pulmonary Arterial Hypertension. Chest 2020;157:376–383. doi: 10.1016/j.chest.2019.09.009

35. Souza R, Badesch DB, Ghofrani HA, et al. Effects of sotatercept on haemodynamics and right heart function: analysis of the STELLAR trial. Eur Respir J 2023;62. doi: 10.1183/13993003.01107-2023

36. Todaro MC, Carerj S, Zito C, et al. Echocardiographic evaluation of right ventricular-arterial coupling in pulmonary hypertension. Am J Cardiovasc Dis 2020;10:272–283. doi:

37. Dardi F, Boucly A, Benza R, et al. Risk stratification and treatment goals in pulmonary arterial hypertension. Eur Respir J 2024;64. doi: 10.1183/13993003.01323-2024

38. Brouckaert J, Verleden SE, Verbelen T, et al. Double-lung versus heart-lung transplantation for precapillary pulmonary arterial hypertension: a 24-year single-center retrospective study. Transpl Int 2019;32:717–729. doi: 10.1111/tri.13409

39. Savale L, Benazzo A, Corris P, et al. Transplantation, bridging, and support technologies in pulmonary hypertension. Eur Respir J 2024;64. doi: 10.1183/13993003.01193-2024

40. Remy-Jardin M, Ryerson CJ, Schiebler ML, et al. Imaging of pulmonary hypertension in adults: a position paper from the Fleischner Society. Eur Respir J 2021;57. doi: 10.1183/13993003.04455-2020

41. Farber HW, Foreman AJ, Miller DP, McGoon MD. REVEAL Registry: correlation of right heart catheterization and echocardiography in patients with pulmonary arterial hypertension. Congest Heart Fail 2011;17:56–64. doi: 10.1111/j.1751-7133.2010.00202.x

42. Fisher MR, Forfia PR, Chamera E, et al. Accuracy of Doppler echocardiography in the hemodynamic assessment of pulmonary hypertension. Am J Respir Crit Care Med 2009;179:615–621. doi: 10.1164/rccm.200811-1691OC

